# Level of Preparedness to Use Psilocybin Among Individuals Seeking Psychedelic Risk Reduction: A retrospective study of a pilot psychiatric consultation service

**DOI:** 10.64898/2026.08.17.26360633

**Authors:** Harland V. Harrison, Mizan Gaillard, Ryan R. Cook, Aryan Sarparast, Ximena A. Levander

**Author notes:** These authors contributed equally to this work as senior authors (Aryan Sarparast, Ximena A. Levander). **Corresponding Author** Ximena A. Levander, MD, MCR, Assistant Professor of Medicine, Oregon Health & Science University, Division of General Internal Medicine & Geriatrics, Addiction Medicine Section, 3181 SW Sam Jackson Park Road Mail Code – UHN24, Portland, OR 97239-3098.

## Abstract

**Introduction:** In 2020, Oregon became the first US state to legalize state-regulated psilocybin services. This study aims to examine: 1) the clinical and demographic characteristics, 2) psilocybin use motivations, and 3) differences in preparedness among patients seeking care in a Oregon-based pilot consult service specializing in psilocybin risk reduction.

**Methods:** This retrospective chart review abstracted sociodemographics, trauma history, and medical and psychiatric risks of patients (November 2023 - September 2025). The Psychedelic Preparedness Scale (PPS), a validated self-report questionnaire, measured preparedness. Two sample t-tests examined associations of PPS scores by insurance, consult motivations, and prior psychedelic use.

**Results:** Patients (N=29) had a mean age of 47.14 years (SD=15.9), were majority female (55.2%); White (82.8%); and privately insured (62.1%). Patients mostly sought psilocybin to address only a psychiatric concern (75.9%); 27.6% anticipated naturalistic (non-state regulated) use. Most patients were deemed low risk for adverse events. Prevalence of prior challenging psychedelic experiences (CPE) was 17.2%; 58.6% reported lifetime psilocybin use. 86.2% endorsed ≥1 form of lifetime trauma. Of PPS completers (N=23, 79%), mean score was 91.3 (SD = 23.99). Scores did not significantly differ by insurance; consultation motivation; CPE; prior psilocybin or psychedelic use.

**Conclusion:** Patients utilizing a novel consultation service demonstrate a high prevalence of trauma, prior psilocybin use, and baseline preparedness. While preliminary, this is among the first descriptions of patients seeking medical and psychiatric consultation when considering psilocybin and highlight the potential role of healthcare systems in providing evidence-based patient education and risk reduction as interest in psychedelics grows.

## Introduction

Recent studies suggest that the use of psilocybin – the active compound in psychedelic mushrooms – is increasing among U.S. adults.^1 2 3 4^ The cross-sectional Global Psychedelic Survey (GPS) conducted in spring of 2023 reported that “psilocybin was used by the highest number of respondents” in the US and Canada.^3^ Between 2004 and 2024, 134 trials were registered to study the use of psilocybin for treatment-resistant depression, major depressive disorder, generalized anxiety disorder, substance use disorders and other health concerns. ^5 6^ Media coverage of these trials has increased significantly since 2020.^7^ This rise of public interest in using psilocybin has likely been fueled by several factors, including state-level decriminalization and legalization and the growing visibility of research on psilocybin as a potential treatment for many psychiatric disorders. Studies have found both an association between decriminalization and increased psilocybin use as well as a higher incidence of mental health disorders among psilocybin users.^8 10^

In November 2020, Oregon voters passed Measure 109, also known as the Oregon Psilocybin Services (OPS) Act, the first U.S. state-regulated legal framework for psilocybin services. OPS created a new avenue for the legal use of psilocybin to paying clients in state-licensed service centers permitted to administer botanical formulations of psilocybin under the supervision of a licensed facilitator, with the first clients seen in 2023.^9^ Psilocybin facilitators must complete a state-approved psilocybin facilitator training program and pass a state licensing exam so they are licensed to provide services – including preparation, dosing, and aftercare, also known as integration.^10^ Between January 1, 2023 to September 30, 2025, there have been 37,963 psilocybin products sold in the Oregon regulated model and 4,577 clients served.^11^ Although many clients report health-related reasons for seeking services, services are not considered to be a medical or clinical treatment, and they do not involve direct medical oversight.^12^ Despite this, the number of emergency response activations and adverse behavioral outcomes in this setting have been minimal.^11^

Because the study of psilocybin, a federally scheduled drug, has been historically restricted in the U.S., our understanding of the short- and long-term effects are still evolving. ^13 14 15 16^ Patients may lack access to consistent and reliable information on the safety of psilocybin use. A 2025 study found a 3-fold increase in psilocybin-related calls at Poison Centers between 2013 and 2022, with most encounters ending with callers seeking in-person treatment at a healthcare facility. ^17^ As the popularity of psilocybin use increases and the legal landscape changes, it is important for medical, mental health, and public health professionals to be better prepared to counsel patients about the potential risks of psilocybin use.^18^

Based in Portland, Oregon, within Oregon Health & Science University (OHSU), the OHSU Psilocybin Education and Assessment Collaborative for Excellence (PEACE) is an outpatient consult service specializing in evaluating risks and counseling patients considering psilocybin use. Patients may present considering use through a state-regulated service center or another setting, which will be referred to as “naturalistic use.” The PEACE clinic model typically includes two approximately 60 to 90-minute appointments. During the first visit, a physician with psychedelic-specific training assesses the patient’s risk and readiness for a psychedelic experience, recording data on patients’ responses in a structured template in their electronic health record (EHR). The second appointment is an educational visit that focuses on risk mitigation. The clinician provides education on the science, physical effects, and use of psilocybin specific to the patients’ health history, risk factors, and needs. Counseling is informed by baseline self-reported preparedness on the Psychedelic Preparedness Scale (PPS), which was developed and applied as a tool to predict outcomes among individuals who go on to ingest psilocybin in a monitored setting.^19^ The clinician will encourage various resources pertinent to the patient’s goals and address questions, which often span themes such as readiness, expectations, medication interactions, subjective effects, and aftercare.

The purpose of this research was to understand the sociodemographics, clinical characteristics, motivation for psilocybin use, and readiness of adults interested in using psilocybin. Our primary aim was to explore the association of various sociodemographic and clinical characteristics on baseline levels of preparedness, as measured by the PPS. Based on preliminary published survey data on US adults,^20^ we hypothesized that patients who are uninsured or on public health insurance, those who intend to use psilocybin to treat a psychiatric disorder, and patients who are high risk, will score lower on the PPS at baseline.

## Methods

### Study Overview

This study was intended as a pilot program where data were abstracted from the EHRs of 29 patients who attended an initial PEACE consult between November 2023 and September 2025. Most consults were completed by a single psychiatrist with subject matter expertise. If consultees met criteria for a substance use disorder within the 12 months preceding the consult, they were seen by an addiction medicine physician also with subject matter expertise. Eligible patients were 18 years and older who completed at least one consult visit within this timeframe. PEACE consultation visits were a mixture of in-person and telehealth, occurred in the state of Oregon, and were billed to the patient’s insurance. This study was approved by OHSU Institutional Review Board (#00029245).

### Data Collection and Procedures

Data were abstracted from the EHRs of eligible patients by 2 researchers using standardized abstraction forms. Data were dual abstracted with discrepancies resolved via discussion between the two researchers, and the larger team, including two clinicians.

### Measures

#### General Demographics

Demographic information included age, race, ethnicity and preferred language. Gender identity was inferred based on information provided by the patient, such as sex assigned at birth and pronouns present in the chart. Information on marriage status, employment, and level of education were gathered from the social history and other descriptive sections taken by the provider in the consult note. Marital status (married or single); employment status (yes/no), and level of education (less than high school, completed high school, associate’s degree or some college, bachelor’s degree, master’s degree, professional degree [e.g. PhD, MD, DO, JD], Other, or Missing). Employment included full time students or those who are self-employed; unemployed included those who receive social security disability payments and who are retired. Primary insurance refers to the main method of insurance at the time of PEACE clinic encounter: private, public (i.e., Medicaid or Medicare), and uninsured.

#### PEACE-Specific Patient Characteristics

The method by which the patient was referred to the clinic (self-presenting; internal or external referral) was collected via the referral documented in the EHR, as well as what was recorded in the encounter note. The structure of PEACE consult notes, which was largely standardized, guided data abstraction for the remaining measures, including variables related to patient social history and measures of past and desired future use of psilocybin and other substances. Diagnoses relevant to the consultation were abstracted from diagnosis coding for the specific PEACE consult encounters.

We used the reason for the referral or the purpose of the consultation as a proxy for what motivated the patient to seek medical consultation prior to psilocybin use and abstracted their reason into five categories 1) addressing a medical concern, 2) addressing a psychiatric concern, 3) medical safety of psilocybin, 4) concerns about psychiatric safety, 5) general mental health, well-being, and religious/spiritual purposes.

During PEACE consults, patients were asked about the lifetime presence of prior sexual, physical, emotional, or medical trauma, and asked about PTSD diagnosis and active or recent PTSD symptoms. Given prior research on associations between trauma and adverse experiences following psychedelic use, ^21 22 23^ history of psychological or physical trauma was also assessed (coded as yes, no, or missing) during PEACE consults. Symptom burden related to trauma history factored into risk ratings.

Information on consult-relevant psychiatric diagnosis were also collected. Unipolar depression included: major depressive disorder (MDD) and persistent depressive disorder (PDD). Anxiety disorders included: generalized anxiety disorder (GAD), panic disorder, and agoraphobia. Obsessive compulsive disorders included: obsessive compulsive disorder (OCD) and mixed obsessional thoughts and acts. Personality disorders included: those clinically suspected after PEACE consult and those that had been previously diagnosed. We left out categories not reported by patients (e.g. schizophrenia).

Medical and psychiatric risks associated with psilocybin ingestion (low, low-moderate, moderate, moderate-high, high) were based in part on literature and judgement of the clinician.^24 25 26 27 28 29 30 31^ While no risk factors were ever removed, some were added as a result of discussion during the consultation (e.g., vascular anomaly). Lastly, no formal personality testing was performed, but personality-related risk factors such as neuroticism were determined based on clinical interview.

#### Information about Prior Psychedelics and Other Substance Use

History of prior psychedelic use was collected from descriptive sections of the PEACE consult note and included: Lifetime use of psilocybin (yes/no), past challenging psychedelic experiences (yes/no), and lifetime use of any psychedelic. We also collected data on what setting (service center or naturalistic use) the patient intended to use psilocybin. Other substance use history included: Past-year substance use (yes/no/missing) including: tobacco, alcohol, cannabis, cocaine, opioids or heroin, and ketamine.

Challenging psychedelic experiences were defined broadly as the patient describing a difficult or distressing experience or impairment during or after the use of a psychedelic.^32 24^ Lifetime psychedelic use was obtained from the patient’s prior psychedelic or non-ordinary states of consciousness experiences and based on whether the patient reported any use of psilocybin, MDMA, LSD, DMT, and ketamine.

#### Preparedness

Level of preparedness to use psilocybin was collected from each patient via The Psychedelic Preparedness Scale (PPS) via Qualtrics.^33^ The PPS is a validated 20-item self-assessment with each item scored on a 7-point Likert scale (1 = not at all, 7 = completely, max total score = 140) that measures different aspects or “factors” of readiness for a psychedelic experience as four subscales.^19 33^ PPS scores for PEACE consults only calculated the total score and did not collect subscale scores. The PEACE PPS also contains the additional item “I don’t understand the question,” coded as a 0, which was added in response to patient feedback about inability to answer certain questions due to inexperience or lack of planning or preparation.

### Statistical Analysis

We obtained descriptive statistics to characterize the variability in the sociodemographics, clinical characteristics, and past substance use history of patients seeking consultation. We evaluated whether baseline PPS score differed based on insurance status, top two consult motivation types, endorsement of challenging psychedelic experience, or reported prior psilocybin or any psychedelic use with Welch’s two-sample t-tests. Spearman’s rank-order correlations were used to determine if there was a correlation between medical risk or psychiatric risk and PPS score. All analyses were conducted in R (v4.5.2), and confidence intervals for Spearman’s correlations were calculated using *DescTools*.^34^

## Results

### Patient Characteristics

A total of 29 patients were included in the sample. The mean age of patients was about 47 years, and the sample was majority female (55.2%), White (82.8%), non-Hispanic/Latino (69.0%), privately insured (62.1%) and held a bachelor’s degree or higher (55.2%) and (**Table 1**). Patients were primarily referred to PEACE internally via the Department of Psychiatry (44.8%) or the Department of Family Medicine (27.6%) and sought to address a psychiatric concern (75.9%) (**Table 1**). A large proportion of patients reported a history of trauma during their PEACE consultation (86.2%) and met criteria for a diagnosis of a depressive disorder and/or PTSD (55.2% and 41.4%, respectively) (**Table 1**).

**Table 1.** General Demographics, PEACE Specific Demographics + Substance Use. General Demographics: Categorical variables are presented as N (%), and numerical variables are presented as Mean (standard deviation). * The total number of patients does not equal 29 due to missing data. For gender, race, and ethnicity, some data are missing because patients declined to provide that information. ^a^ If patients listed more than one race, they were placed in this category alone (and thus not double counted). PEACE Specific Demographics: ^a^ If patients listed more than one motivation, they were placed in this category alone (and thus not double counted). * The total number of patients does not equal 29 due to missing data. Psychedelics and Other Substance Use: ^a^ Inclusive of psilocybin Percentages are calculated based on a denominator of 29 to reflect the percentage of PEACE patients that reported having used that substance in the past year.

| General |  |
| --- | --- |
|  | Value |
| Age (Years) | 47.14<br>(15.9) |
| **Gender\*** |  |
| Female | 16 (55.2) |
| Male | 11 (37.9) |
| Gender Diverse | 1 (3.5) |
| **Race\*** |  |
| White | 24 (82.8) |
| Two or More<sup>a</sup> | 1 (3.5) |
| **Ethnicity\*** |  |
| Hispanic/Latino | 2 (6.9) |
| Not Hispanic/Latino | 20 (69.0) |
| **Marriage/Partnership Status\*** |  |
| Yes | 15 (51.7) |
| No | 12 (41.4) |
| **Employment\*** |  |
| Yes | 17 (60.7) |
| No | 10 (35.7) |
| **Highest Education Level\*** |  |
| High School | 1 (3.5) |
| Associate's or Some College | 5 (17.2) |
| Bachelor's | 8 (27.6) |
| Master's | 6 (20.7) |
| Professional | 2 (6.9) |
| **Primary Insurance** |  |
| Private | 18 (62.1) |
| Public | 11 (37.9) |
| **PEACE** |  |
|  | **N (%)** |
| **Referral Method** |  |
| Department of Psychiatry | 13 (44.8) |
| Department of Family Medicine | 8 (27.6) |
| Self-presenting | 6 (20.7) |
| External | 2 (6.9) |
| **Consult Motivation** |  |
| Psychiatric Concern | 22 (75.9) |
| Two or More Motivations <sup>a</sup> | 4 (13.8) |
| General Mental Health and Wellbeing | 1 (3.5) |
| Psilocybin Interaction with Psychiatric Concern | 1 (3.5) |
| Medical Concern | 1 (3.5) |
| **Anticipated Use Setting\*** |  |
| Service Center | 12 (41.4) |
| Naturalistic Use | 8 (27.6) |
| **Reported Trauma History** |  |
| Yes | 25 (86.2) |
| No | 4 (13.8) |
| **Consult-Relevant Psychiatric Diagnoses** |  |
| Unipolar Depression | 16 (55.2) |
| Posttraumatic Stress Disorder (PTSD) | 12 (41.4) |
| Anxiety Disorder | 10 (34.5) |
| Substance Use Disorder | 4 (13.8) |
| Obsessive Compulsive Disorders | 3 (10.3) |
| Bipolar Disorder | 2 (6.9) |
| Personality Disorder | 2 (6.9) |
| Eating Disorder | 1 (3.5) |
| Functional Neurologic Disorder | 1 (3.5) |
| **Psychedelics and Other Substance Use** |  |
|  | **N (%)** |
| **Lifetime Use of Any Psychedelic <sup>a</sup>** |  |
| Yes | 21 (72.4) |
| No | 8 (27.6) |
| **Challenging Psychedelic Experience** |  |
| Yes | 5 (17.2) |
| No | 22 (75.9) |
| **Lifetime Use of Psilocybin** |  |
| Yes | 17 (58.6) |
| No | 12 (41.4) |
| **Past-Year Substance Use<sup>b</sup>** |  |
| No | 9 (31.0) |
| Yes | 20 (69.0) |
| Cannabis | 14 (48.3) |
| Alcohol | 13 (44.8) |
| Tobacco | 3 (10.3) |
| Ketamine | 2 (6.9) |
| Opioids or Heroin | 1 (3.5) |

While many patients reported lifetime use of a psychedelic (72.4%), with approximately 58% citing use of psilocybin specifically, fewer endorsed having a challenging psychedelic experience (17.2%), (**Table 1**). Several patients also endorsed past-year substance use (69.0%), with cannabis (48.3%) and alcohol use predominating (44.8%) (**Table 1**).

#### PPS Scores

Results revealed that for those who completed the PPS (n =23, 79%), their mean score was relatively high at 91.3 (SD = 24.0, 95% CI [80.9, 101.7]) (**Figure 1**). All Welch’s two-sample independent t-tests, performed to explore whether PPS score differed based on characteristics and psychedelic use-related factors, revealed null results (**Table 2**). However, the difference in scores between patients who reported prior psilocybin use (M = 101.0) and those who did not (M = 82.3) trended towards significance, (p = 0.06). Similarly, the difference in scores between those who reported no prior psychedelic use at all (M = 85.3) and those who reported using psychedelics one or more times (M = 110.0) also trended towards significance (p = 0.06)

**Table 2.** Mean Psychedelic Preparedness (PPS) Scores by Demographic Characteristic P-values are the result of Welch’s two-sample independent t-tests exploring whether PPS score differed based on demographic characteristics.

|  | Mean PPS Score (SD, N) | p-value (95% CI) |
| --- | --- | --- |
| Primary Insurance Type |  |  |
| Private | 90.0 (26.1, 16) | 0.68 (-25.4,17.1) |
| Public | 94.1 (20.0, 7) |  |
| Consult Motivation |  |  |
| Psychiatric Concern | 92.2 (25.4, 19) | 0.69 (-39.9,52.2) |
| Multiple | 86.0 (22.3, 3) |  |
| Challenging Psychedelic Experience |  |  |
| Yes | 95.8 (29.9, 4) | 0.75 (-50.1,39.1) |
| No | 90.3 (24.8, 17) |  |
| Prior Psilocybin Use |  |  |
| Yes | 101.0 (21.7, 11) | 0.06 (-38.2, 0.84) |
| No | 82.3 (23.3, 12) |  |
| Prior Psychedelic Use |  |  |
| Yes | 110.0 (19.7, 7) | 0.06 (-50.7,1.26) |
| No | 85.3 (16.5, 4) |  |

**Figure 1.**
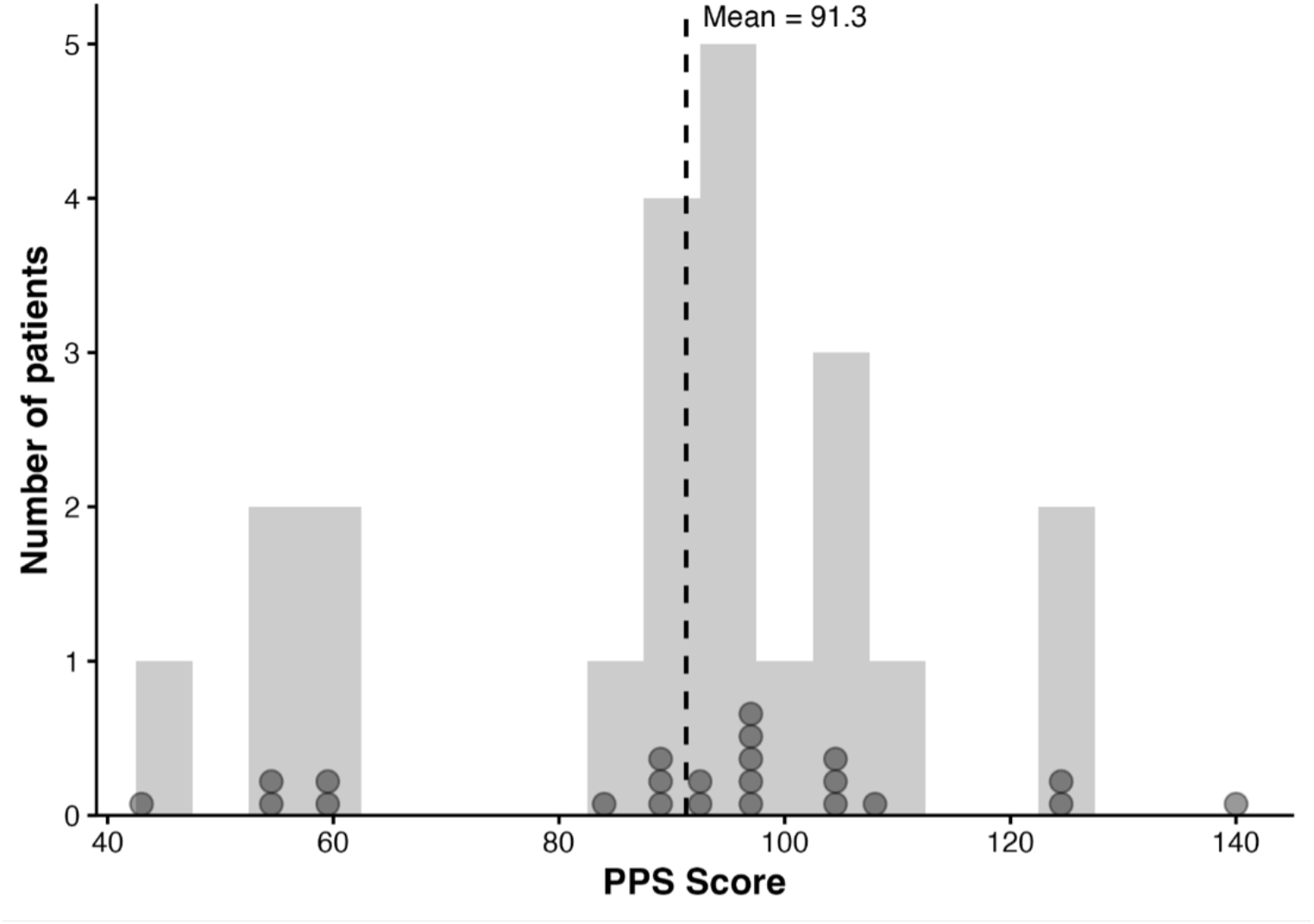
Psychedelic Preparedness Scale (PPS) Score Distribution Distribution of psychedelic preparedness scale (PPS) scores, with a vertical broken line denoting the mean and dots showing the exact score of each participant with available data (n = 23).

The results for Spearman’s rank-order correlations between each risk rating (**Table 3**) and PPS scores were also non-significant. For the medical risk correlation, ρ = −0.07, n = 23, p = 0.74, 95%CI [−0.47, 0.35], and for the psychiatric risk correlation, ρ = −0.09, n = 23, p = 0.69, 95% CI [−0.48, 0.34].

**Table 3.** Medical and Psychiatric Risk. Risk assessments were assigned based on clinical impression and literature detailing risk factors for adverse events related to psilocybin use. See supplementary material for further details. Some patients did not receive a risk rating and thus the n reported here does not sum to 29 for either risk type.

| Rating | N (%) |
| --- | --- |
| **Psychiatric Risk** |  |
| Low | 11 (37.9) |
| Low to Medium | 3 (10.3) |
| Med | 8 (27.6) |
| Medium to High | 2 (6.9) |
| High | 2 (6.9) |
| **Medical Risk** |  |
| Low | 16 (55.2) |
| Low to Medium | 0 |
| Med | 9 (31.0) |
| Medium to High | 1 (3.5) |

## Discussion

This pilot study outlines a novel model of healthcare response to growing public interest in psilocybin use and a rapidly evolving legal landscape – an outpatient consult clinic focused on risk reduction and counseling patients housed within an academic medical center. Overall, our results demonstrate that those seeking medical advice prior to psilocybin use through the PEACE clinic have a generally high sociodemographic status (i.e., employed, privately insured, partnered, highly educated),^35^ have common psychiatric disorders studied or being studied by current psychedelic clinical trials, have low rates of prior challenging psychedelic experiences despite high rates of prior psychedelic use, and are generally assessed at a low level of risk for adverse events. This study population also had a high level of preparedness before being provided any risk counseling, and it appears preparedness at the time of consult may not be strongly related to markers of socioeconomic status, consult motivation, or prior experiences with psychedelics. Thus, the PEACE clinic appears to be a viable method of providing much-needed risk reduction and patient education services to a subset of the growing number of people seeking to use psilocybin outside of research settings.

Notably, 86% of patients reported experiencing some form of trauma in their lifetime, which is higher than the estimated 62.8% of US adults who had past exposure to an adverse childhood experience (ACEs), which are often traumatogenic, or the 70% of adults worldwide who report having experienced at least one lifetime traumatic event.^36^ To date, there have been no published clinical trials on psilocybin as a therapy for PTSD, although it appears to demonstrate potential as a therapeutic agent.^37^ The OPS 303 Client Data Form, which collects demographic and motivation for use data from OPS clients, asks about multiple forms of trauma, and recent published data from the OPS Data Dashboard from Q1-Q3 2025 data shows that many clients cite trauma and mental health disorders as a reason for seeking OPS services.^11^ These data align with the motivations for seeking treatment among our patients and may contribute to the main reason for consultation being a psychiatric rather than medical concern. Our results suggest that patients may be seeking to use psilocybin to help mitigate mood and anxiety symptoms as well as symptoms of trauma related disorders. Given public interest in psilocybin for treatment of trauma-related disorders, more research is needed to understand how psilocybin may impact trauma-related outcomes.

Although PEACE is housed within a tertiary care medical center that serves a medically complex patient population, the patients included in the present study were assessed as having a generally low level of risk for experiencing an adverse event related to psilocybin use. The limitations of a harm-reduction model of care in favor for a risk management approach are thus highlighted when considering the low risk of harm among this population and the possibility of improved mood and anxiety disorder symptoms shown in clinical trial data. ^38 39 6^ Greater emphasis could be placed on counseling patients about taking steps to create a physical and psychological environment that maximizes the potential benefits of psilocybin use, normalizing challenging experiences, and encouraging patients to arrange follow up aftercare appointments with a trusted peer or professional after a psilocybin experience. These actions have been shown to increase positive outcomes overall and may warrant recharacterizing similar consultation services as risk mitigation or wrap-around care, rather than harm-reduction, which is applied to substances associated with a much higher probability of harmful outcomes.^40 41 42 43^

Interestingly, the majority of our study sample were highly educated, well-resourced individuals, 75% of whom expressed interest in using psilocybin in a service center, which can cost hundreds to thousands of dollars. They also had a high mean score on the PPS, more than half reported prior psilocybin use, and an even greater portion demonstrated experience with at least one other psychedelic. It is curious that patients with substantial means and prior psychedelic experiences are seeking care at the PEACE clinic, which could be influenced by a commonality among psychedelic enthusiasts and the novelty of the consult service center model as motivators for these patients to seek consultation, or historically inequitable access to psilocybin and education about psychedelics in the US^44^. A better understanding of why well-resourced and well-informed patients sought out psychedelic-related consultation warrants further research. It could indicate broader clinician discomfort with discussing risks related to psilocybin use (hence referring patients for consultation). Given the large variation of prior psychedelic use among clinical trial participants^45^ characterizing and orienting under-resourced and psychedelic-naïve individuals is an important research topic in addressing barriers to accessing psychedelic education and examining the relationship between risk and psychedelic literacy.

Somewhat surprisingly, average PPS score did not vary as a function of any of the measures tested, including prior psychedelic use and challenging psychedelic experiences, and neither risk type was correlated with PPS. While analyses using insurance type, consult motivation, and CPE were far from statistically significant, those testing the relationship between PPS and use of any psychedelic and prior psilocybin use both trended towards significance (p = 0.06). The relationship between risk level and preparedness may also be different for similar reasons. Studies with larger samples are needed to clarify which patient characteristics are most related to level of preparedness and how counseling correlates with meaningful PPS score changes.

### Limitations

The PPS was initially developed as a research tool rather than to guide clinical care or psychoeducation, so its clinical utility is still being investigated and optimized. Of the 29 PEACE consult patients, only 23 of them had PPS score data and this was obtained by patient visit documentation which could be prone to error compared to direct collation of scores. PEACE is a small clinic that does not have full time staff, and the PPS was administered as part of clinical care rather than as a research tool. Thus, no reminders to complete it were sent, and it was not necessary for consults. The small sample sizes for this pilot (n = 29) and PPS-related analyses (n = 23) both limit the generalizability of our findings and power to detect effects of interest; however, the demographics reported by the most updated OPS data at time of publication (aggregating data from January 2025 through September 2025) are similar to those of this investigation. About 77% of OPS clients identified as White (with 17% preferring not to answer questions about racial identity) and almost 54% were women. Thus, while our sample sizes are small, they are demographically similar to the most recent data tracking those utilizing the state-regulated psilocybin services model. Second, due to patient feedback, an extra response, “I don’t know”, was added to the PPS as it was used in PEACE. While the addition of this extra question may have compromised the psychometric validity of the PPS, it allowed PEACE clinicians to be responsive to patient needs, a core goal of the consult service. Lastly, the PPS has subscales, which could not be calculated for the present analyses. We thus used only the summary PPS score, which limited our ability to draw conclusions about how patient characteristics related to specific dimensions of preparedness. Future research should leverage both the summary and subscale scores of the PPS. Future research could examine what patients are desiring in psilocybin education to guide counseling session and could follow patients after psilocybin administration session(s) to examine how psilocybin education may have impacted their experiences.

## Conclusion

This retrospective study of a pilot psilocybin consultative clinic provides one of the first descriptions of patients seeking medical and psychiatric consultation when considering psilocybin use. It focuses on risk assessment, patient education, and risk reduction rather than treatment outcomes from psilocybin use, which addresses a critical but under-studied dimension of psychedelic public health. By informing clinicians and public health practitioners, about the characteristics and needs of individuals seeking psilocybin-related services, this work contributes to developing evidence-guided risk reduction approaches and supporting informed decision making. As psychedelics become more popular and accessible this work contributes to ongoing discussions about how public health and healthcare systems can responsibly respond to increasing psychedelic use outside formal clinical trials.

## Data Availability

All data produced in the present study are available upon reasonable request to the author

## Author Contributions

HVH (conceptualization, investigation, methodology, data curation, writing, review & editing, project administration, funding acquisition), MG (data curation, methodology, formal analysis, writing, review & editing), RRC (supervision, review & editing), AS (conceptualization, supervision, writing, review & editing), XAL (conceptualization, supervision, writing, review & editing)

## Funding

This research was made possible by a grant awarded through the N.L. Tartar Trust Research Fellowship (2025-2026) awarded through the Oregon Health & Science University Foundation. The content of this manuscript and research is solely the responsibility of the authors and does not represent the official views of funders and funders were not involved in the decision to publish findings or in the writing process.

## Conflicts of Interest

AS is the founder of Penumbra Consultation, a private practice providing psychedelic-related consultation to patients and healthcare providers. Thie entity had no role in the design, conduct, analysis, interpretation, or funding of this study, or in the preparation of the manuscript.

The remaining authors declare that the research was conducted in the absence of any commercial or financial relationships that could be construed as a potential conflict of interest.

## Notes

### Author Declarations

Institutional Review Board of Oregon Health & Science University waived ethical approval for this work.

