## Supplemental Materials for "Level of Preparedness to Use Psilocybin Among Individuals Seeking Psychedelic Risk Reduction: A retrospective study of a pilot psychiatric consultation service"

Supplemental Material

Psilocybin Risk Assessment Preface:

Reviewing all clinical trials (n = 11) on psilocybin from 1991-2021, summing a review of 257 individuals, common side effects of psilocybin are: nausea, fatigue, headache, hypertension, dysphoria (anxiety, sadness, etc). Less common (< 2-20%) was tachycardia, ataxia, transient psychotic symptoms (namely paranoia). Note that 40% of participants with depression enrolled in clinical trials involved feelings of panic, and 31 - 39% of even healthy individuals without mental health conditions experienced strong or extreme fear, though there was no correlation found between degree of dysphoria and degree of benefit.^1^ ^2^ ^3^ These adverse effects resolved typically within 24h. Limiting the data are small sample sizes (often <50) with a preponderance of White, college-educated individuals, often with prior psychedelic experiences. Excluded from trials were individuals with significant underlying cardiovascular, neurologic, hepatic, or renal conditions. Trials utilized controlled settings with well-screened participants including a range of 12 - 35 hours of combined prep, supervision, and aftercare with masters-level therapists or beyond.^4^ ^5^ Therefore, the medical risk factors compiled below are based on the aforementioned information, recognizing the absence of data as a risk factor.

Simonsson et al 2023 identified 6 set & setting variables that act as potential predictors of a challenging psychedelic experience.^6^ Studerus et al 2012 identified 3 additional risk factors that increase the risk of an anxious response to a psychedelic experience.^7^ Psilocybin also carries a risk of precipitating psychosis and mania among those predisposed for these conditions such as a personal symptoms or that of a first degree relative, therefore these individuals have been excluded for modern psychedelic clinical trials. In combining the available literature on risk factors for negative psychological outcomes, we have listed the applicable major risk factors below.

**Psilocybin Medical Risk Assessment:**

The following risk factors increase medical risk from psilocybin ingestion. Primary concerning adverse outcomes medically are cardiac events given psilocybin's pressor effects on blood pressure and heart rate. Note that risk factors are based on available data (see preface above) and available clinical expertise.

*Age > 65*

*History of syncope*

*History of persistently low (<90/60) or high blood pressure (>140/90)*

*History of heart palpitations or arrhythmia*

*History of heart attack*

*Elevated QT interval if known*

*First degree relative with Premature Heart Disease (I.e. heart attack or stroke before the age of 60)*

*History of Congenital Heart Disease or Structural Heart Disease*

*History of Heart Failure*

*History of Of Atherosclerosis*

*History of Hyperlipidemia*

*History of Obestiy*

*History of Congenital Exposure to Rubella*

*History of Rheumatic Fever*

*History of Endocarditis*

*History of Carcinoid Tumor*

*History of Autoimmune Disease (e.g. Lupus, rheumatoid arthritis)*

*History of Smoking Tobacco*

*History of Gastrointestinal Disease (e.g. cyclical vomiting syndrome, inflammatory bowel disease, cyclical vomiting syndrome, diabetic gastroparesis)*

*History of Traumatic Brain Injury*

*History of Seizures/Epilepsy*

*History of Allergic response to medicinal or culinary mushrooms or psilocybe mushrooms*

*History of Severe Asthma*

**Psilocybin Psychological Risk Factors:**

The following risk factors increase psychological risk from psilocybin ingestion*.* Primary concerning adverse psychological outcomes are traumatic or challenging psychedelic experiences, psychosis, mania, or exacerbation of underlying mental health condition(s). Risk factors are based on available data (see preface above) and available clinical expertise.

*Intent for use in an uncontrolled setting*

*High active trauma-related symptoms burden*

*Limited or absent support network*

*Limited or absent trained psychotherapy support*

*Intent for use while alone without support person present*

*Low score on PPS measure, if completed*

*No plan to determine dosage of psilocybin*

*Plan to use high dosage of psilocybin*

*Absence of prior psychedelic experiences*

*Disagreeable social or physical setting*

*High levels of neuroticism*

*High levels of emotional excitability*

*Low levels of absorption*

*Adolescence and/or young age*

*Co-use of lithium or haloperidol*

*Recent major life event*

*Personal history of significant psychosis symptoms*

*Personal history of confirmed bipolar disorder*

*Personal history of borderline personality disorder*

*First or second degree relative with a psychotic disorder*

*First or second degree relative with a bipolar disorder*
